# Intracranial Photobiomodulation In De Novo Parkinson’s Patients: preliminary data of an ongoing randomized controlled trial

**DOI:** 10.1101/2025.01.23.25320819

**Authors:** Stéphan Chabardès, Stéphane Thobois, John Mitrofanis, Maude Beaudoin-Gobert, Daniel Anglade, Nicolas Aubert, Vincent Auboiroux, Pierre Bleuet, Thomas Costecalde, Emmanuel De Schlichting, Olivier Faivre, Marine Mastrosimone, Jenny Molet, Adrien Poizat, Sarah Renault, Napoleon Torres, Claude Chabrol, Cecile Moro, Alim Louis Benabid

## Abstract

Parkinson’s disease (PD) is a progressive neurodegenerative disorder and current treatment options only tackle symptoms. We aimed to evaluate the effect of intracranial photobiomodulation (iPBM) on dopaminergic neurons and its consequences on motor symptoms in de novo PD patients.

This randomised open label study included 6 de novo PD patients, 3 served as control and 3 were treated by chronic iPBM during 2 years. Researcher analysing the PET data was unaware of patients’ iPBM status but clinical examinator was aware of it. Data analysis was performed on June 2024.

De novo PD patients were recruited and randomized (1:1) to receive iPBM or medical treatment alone for 4 years. PD diagnosis was < 2 years, and no medication was allowed at inclusion. PET scan was performed at baseline and at 1 year

Primary outcome was safety. The secondary outcome measure was the evolution at 2 years of motor scales using the MDS-UPDRS part III and the evolution of PET scan (using the [11C]-PE2I ligand) at 1 year.

Six patients (1f, 5m) were recruited with a mean age (SD) of 55.3 years (7.6). We showed that iPBM was feasible and safe on the first 3 operated patients. The first novelty of the study was that iPBM improved motor scores in average at 2 years whereas the control group followed the natural clinical decline. Mean (SD) clinical outcome with iPBM was -6.2% (32.9%) improvement on the MDS-UPDRS (part III) compared to baseline versus +130% (143%) for control group. The second novelty was that in the iPBM group, [11C]-PE2I PET scans showed an unusual mean increase in tracer binding within the caudate +21.3%(15.9), nucleus accumbens +22.2%(23.7%), pallidum +89.7% (94.8%) and putamen +26.1%(34%). in the control group, as expected, tracer binding was decreased (SD) on average in the caudate - 16.9%(−16.3%), nucleus accumbens -27.5%(−28.4%), and putamen -19.5%(−11.4%), but slightly increased in the pallidum +10.7%(+39.6%). Because of the small number of patients, no statistical test was performed at this stage.

**Conclusions:** iPBM was feasible and safe in this small group of patients. It showed an unusual improvement of motor scores at 2 years that was associated with striking increase of dopamine transporter in the basal ganglia observed on the [11C]-PE2I PET. These preliminary data suggest a possible modifying disease effect related to iBPM. A larger number of patients with longer follow up is mandatory to validate our preliminary results.

**Trial Registration:** the EvNIR study is registered at ClinicalTrials.gov (NCT04261569).

## INTRODUCTION

Parkinson’s disease (PD) is a progressive neurodegenerative disorder affects 1 to 2% of over 60-year-olds population. Current treatment options only tackle symptoms, and there is no way to forestall or slow down the neurodegenerative process. Despite extensive research, no therapy has yet shown disease-modifying capacities ^1–3^. However, a very recent study suggested that Lixisenatide, a glucagon-like peptide 1 receptor agonist, could slow the progression of motor disability ^4^. An effective disease-modifying treatment remains a major unmet need for PD patients to delay progression of their severe irreversible motor and non-motor disabilities.

Mitochondrial dysfunction is one of the key mechanisms leading to the loss of dopaminergic neurons located in the susbtancia nigra compacta (SNc) ^5,6^. Indeed, it is well known that: 1) MPP+, a metabolite of MPTP, inhibits mitochondrial complex I, inducing a specific degeneration of SNc neurons; 2) Rotenone, a pesticide that can induce PD, acts by inhibiting mitochondrial complex I in SNc neurons; 3) Pathogenic variants of the genes *PINK1* (encoding PTEN-induced putative kinase 1), *Parkin* or *DJ1* are associated with deficits in mitochondrial function ^6–8^.

Photobiomodulation (PBM) – i.e., the stimulation of tissues using red to near infrared light (λ=600-1300 nm) – can restore mitochondrial function, resulting in a short-term increase in adenosine triphosphate (ATP) energy production in cells and the expression of stimulatory and protective genes^9^. We have already shown that PBM can enhance survival of dopaminergic neurons and improve motor behaviour^10–12^.

The aims of the study are to assess safety, motor evolution as well as dopaminergic denervation after chronic iPBM. Herein we report on the first 3 treated patients compared to 3 control PD patients.

## METHODS

### Patients

This study was conducted between 2020 and 2024, it is part of an ongoing randomised, open-label, case-control study that will ultimately include 14 patients. The first six consecutive patients *(see table 1)* are reported here. They met all the following inclusion criteria: PD diagnosis within the last 2 years, naïve of any anti-PD drugs at the time of inclusion, Hoehn and Yard stage 1 or 2, and dopamine denervation confirmed on baseline PET and [^11^C]-PE2I, a dopamine transporter ligand ([^11^C]-PE2I-PET). After inclusion, patients were allowed to take anti-parkinsonian drugs if required except Rasagiline.

**Table 1.** General characteristics of patients and clinical data. ^*^: Control #1 declined to take anti parkinsonian medications despite the medical advice given by his neurologist. LEDD: Levodopa Equivalent Daily Dose was computed based on Tomlison C et al^19^. MDS-UPDRS: Movement Disorder Society-Unified Parkinson Disease Rating Scale MDS-UPDRS scores part 3 for the three Control patients worsened over time, whereas scores for iPBM patients iPBM#1 and #2 improved. For iPBM #3, after initial worsening at 12 months follow-up, scores showed an improving trend, following the adjustment of the infrared light exposure schema.

|  | Pbm 1 | Pbm2 | Pbm3 | Control 1 | Control 2 | Control 3 |
| --- | --- | --- | --- | --- | --- | --- |
| **Age at inclusion**<br>(years) | 60's | 50's | 60's | 60's | 60's | 40's |
| **Randomisation** | PBM | PBM | PBM | Control | Control | Control |
| **Time since diagnosis**<br>(months) | 16 | 25 | 27 | 14 | 17 | 19 |
| **Hoehn and Yahr**<br>stage<br>at baseline | 2 | 2 | 2 | 2 | 2 | 2 |
| **History of PD**<br>(personal/familial) | no | no | no | no | no | no |
| **Symptoms at onset** | Left upper limb, rigidity, akinesia micrographia | Left upper limb rigidity, akinesia dysarthria | Left upper limb rigidity, akinesia | Right upper limb akinesia tremor micrographia | Right upper limb tremor | Left lower limb tremor, rigidity |
| **LEDD (mg/ DAY)** |  |  |  |  |  |  |
| Baseline | 0 | 0 | 0 | 0 | 0 | 0 |
| M6 | 300 | 255 | 0 | 0* | 405 | 303 |
| M12 | 450 | 202 | 0 | 0* | 457 | 424 |
| M18 | 450 | 377 | 80 | 0* | 607 | 764 |
| M24 | 450 | 402 | 460 | 0* | 733 | 1114 |
| **MDS-UPDRS**<br>part III |  |  |  |  |  |  |
| Baseline | 38 | 18 | 19 | 20 | 29 | 17 |
| M6 | 38 | 15 | 33 | 20 | 24 | 40 |
| M12 | 34 | 21 | 40 | 25 | 31 | 52 |
| M18 | 23 | 20 | 39 | 29 | 32 | 57 |
| M24 | 23 | 17 | 24 | 33 | 38 | 67 |

All patients gave their written informed consent, and the clinical trial was approved by the French National Agency for the Safety of Medicines and Health Products (ANSM) in January 2020, and by the local ethics committee in April 2020. The study is registered at ClinicalTrials.gov (NCT04261569).

### Pre- and post-surgery assessments

Motor signs were assessed every 6 months by the same examinator using the Movement Disorders Society-Unified Parkinson Disease Rating Scale (MDS-UPDRS), part III, without treatment at baseline, and after 24 hours of anti-Parkinson drug withdrawal for later assessments. Walking analysis was performed at baseline and at 12 months using Vicon software on a dedicated platform.

Eye fundus was examined by an independent ophthalmologist at baseline and every 6 months thereafter.

**S**urgery, laser probe and illumination settings are presented in figure 3.

## RESULTS

Patients control#1, #2 and #3 did not receive any iPBM device. The 3 patients randomised to the iPBM group (PBM#1, #2 and #3) received iPBM that was turned ON five days after surgery.

### Clinical evaluation

(See table 1 and figure 1).

**Figure 1:**
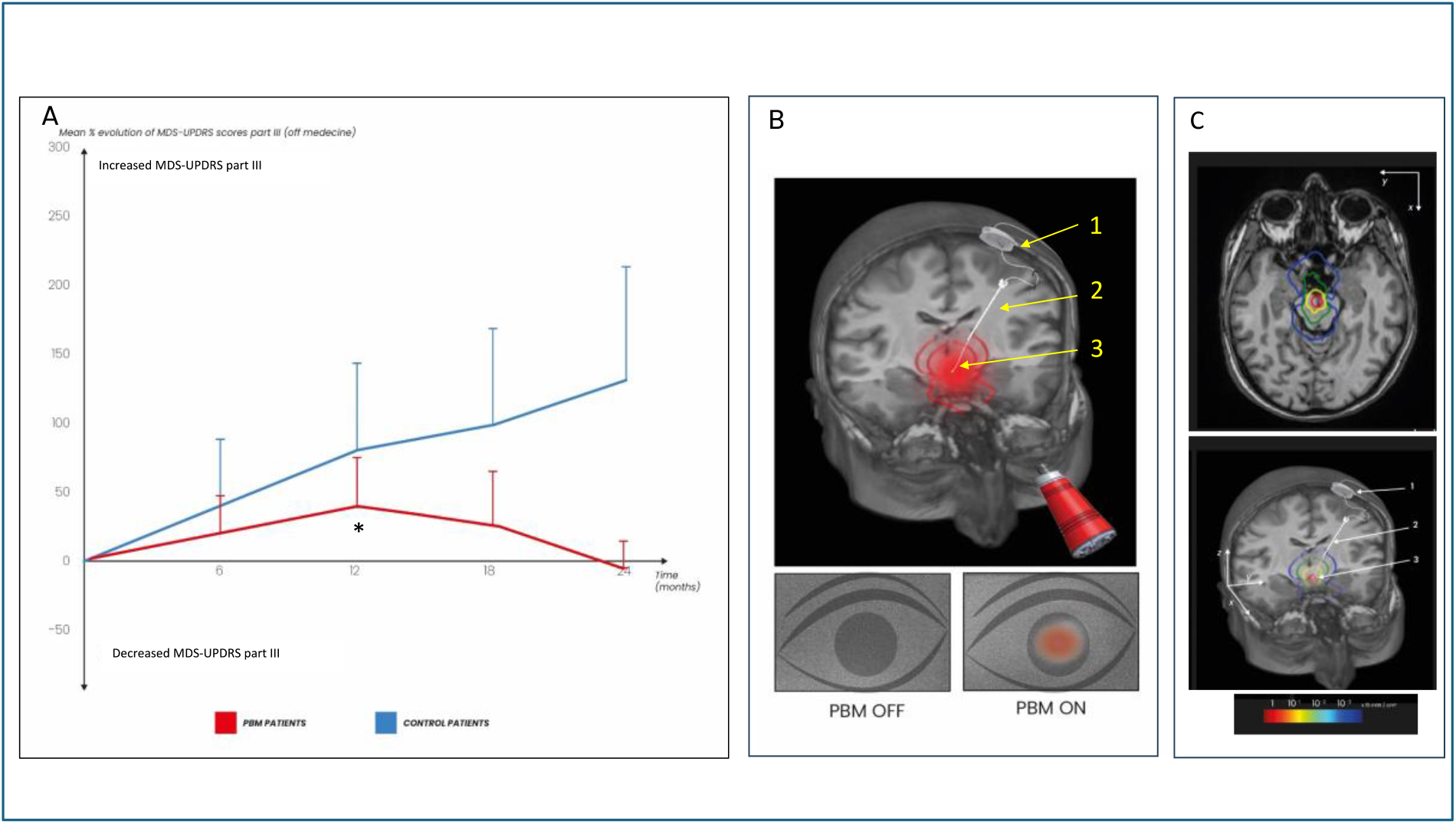
Evolution of mean MDS-UPDRS score part III and PBM device. iPBM group: patients iPBM#1, #2, #3; Control group: patients Control #1, #2, #3: A: Group analysis of the percentage evolution of MDS-UPDRS motor scores part III for iPBM group and controls patients between baseline and 24 months, after 24 h OFF med conditions. Positive values represent worsening symptoms. Individual values can be found in Table 1. *: at M12, infrared light exposure duration was changed from 1 minute ON/ 4 minutes OFF 24 h/day to 2 minutes ON/ 2 minutes OFF 24 h/day in iPBM #3. B: Intra-cranial device for iPBM and simulation of photons propagation in the cranium superimposed to patient #1 MRI and camera detection in patient #1. (see online-only material). Device for iPBM, consisting of a laser box (1) inserted into the skull, a ventricular catheter (2) used to guide the laser probe (3) into the lateral ventricle and toward the third ventricle. In the 20-s exposure images recorded by the camera, when the laser is off and on, a clear spot is observed when the laser is switched on: the eye is backside-illuminated and most of the light naturally exists through the pupil (brightest spot).

Mean (SD) MDS-UPDRS part III scores (maxi 132) OFF-medication improved from 25 (11) at baseline to 21 (3) at 24 months in the iPBM group versus 22 (6) at baseline to 46 (18) at 24 months in the control group.

Indeed, a substantial improvement or stabilisation of MDS-UPDRS part III scores was observed for iPBM#1and iPBM#2 at 24 months. For iPBM#3, motor scores worsened from 19 to 40 at 12 months follow-up. At this stage, we increased the daily duration of illumination (2 minutes ON and 2 minutes OFF, compared to the initial cycle of 1 minute ON and 4 minutes OFF), without modifying the power. Following this adjustment, MDS-UPDRS part III initially plateaued, reaching 39 at 18 months, and subsequently improved, reaching 24 at 24 months.

In contrast, in the control group, MDS-UPDRS part III scores deteriorated over time, as expected in PD.

Furthermore, looking at gait, walking speed, cadence and step lengths improved for the iPBM group on average at 12 and 24 months (see online-only material and eFigure2 in supplement 3). In the Control Group, cadence slightly improved on average but step lengths and walking speed slightly deteriorated.

### Medications: (see table 1)

Mean Levodopa Equivalent Daily Dose (LEDD) at 24 months (SD) was 437 mg (31) for patients in the iPBM group. In control group, mean LEDD at 24 months was 615 mg (566).

### PET Imaging (see figure 2)

#### iPBM Group

[^11^ C]-PE2I-PET at 12 months compared to baseline showed a marked mean (SD) bilateral increase of [^11^C]PE2I BPND in the caudate nucleus +21.3% (15.9), the nucleus accumbens +22.2% (23.7%), the putamen +26.1% (34%), and the pallidum +89.7% (94.8%).

**Figure 2:**
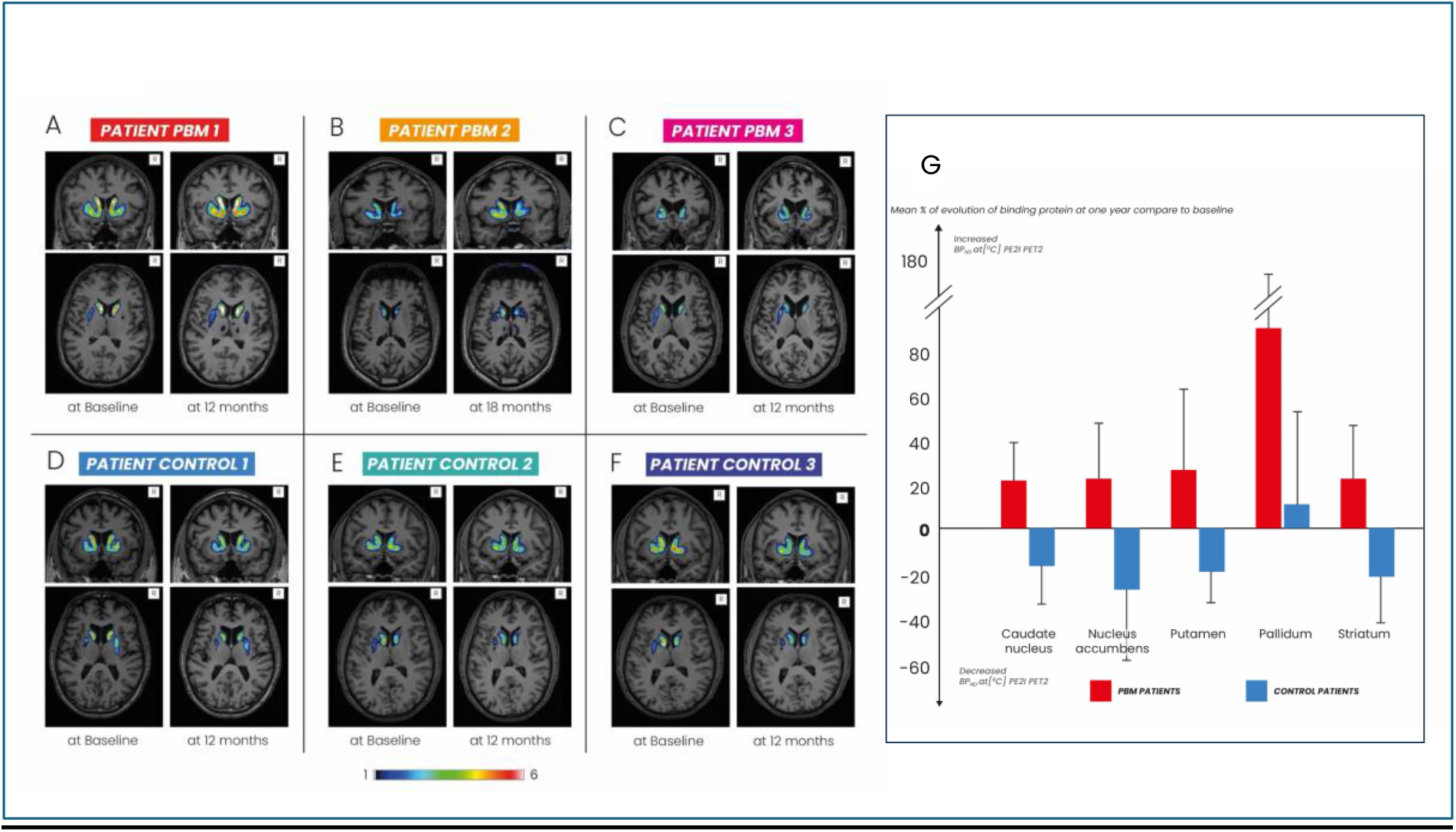
Evolution of [^11^C]-PE2I binding. A to F: presynaptic dopaminergic denervation as determined from [^11^C]-PE2I PET scans at baseline and at 12 months in iPBM group and at 12 months in Control group. A: In iPBM #1 at baseline, a bilateral reduction of [^11^C]-PE2I binding predominates in the putamen, mostly on the right side. Bilateral increase of tracer binding in the putamen and caudate is observed at 12 months. B: In iPBM #2, [^11^C]-PE2I binding is reduced on both putamen and caudate at baseline. At 12 months, increased tracer uptake is observed bilaterally in the caudate and putamen. C: In iPBM #3, [^11^C]-PE2I binding is reduced on both putamen and caudate at baseline. A slight decrease in tracer binding is observed bilaterally in the putamen and caudate at 12 months. D to F: In controls #4, #5 and #6, overall reduction of striatal [^11^C]-PE2I is observed at baseline, with worsening at 12 months follow-up. G: Variations in bilateral non-displaceable [^11^C]-PE2I binding potential (BPND) between baseline and 12 months follow-up at group level. BP_ND_ was measured in the caudate nucleus, nucleus accumbens, putamen, and pallidum in all patients. iPBM group showed a remarkable BP_ND_ increase in all structures. Control patients showed BP_ND_ decrease in all structures except the pallidum (mainly in control #2).

**Figure 3:**
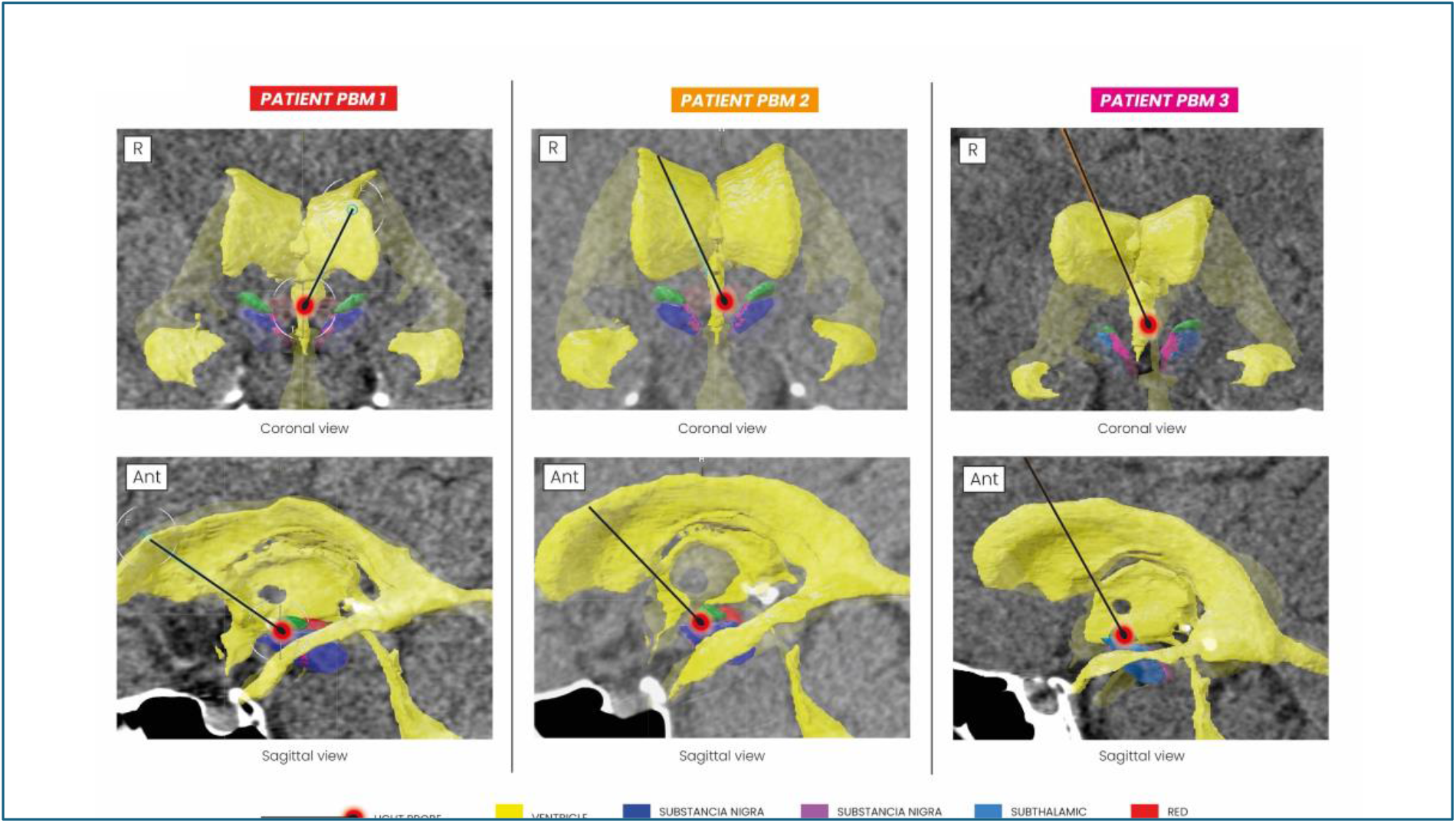
Localization of laser probe. The laser probes were inserted unilaterally from the left or right side depending on vein anatomy and ventricle constraints. The trajectory of the probe followed the lateral ventricle, the foramen of Monro and finally the 3^rd^ ventricle as already described^18^. The tip of the laser probe was intended to be localized on the floor of the third ventricle, on the midline, approximately 6 to 7 mm from the medial edge of the SNc. Mean coordinates (relative to AC-PC line and Posterior Commissure) of the tip of the laser probe were as follows: x = +11.6; y = -0.9; z = -5.2.

**Figure 4:**
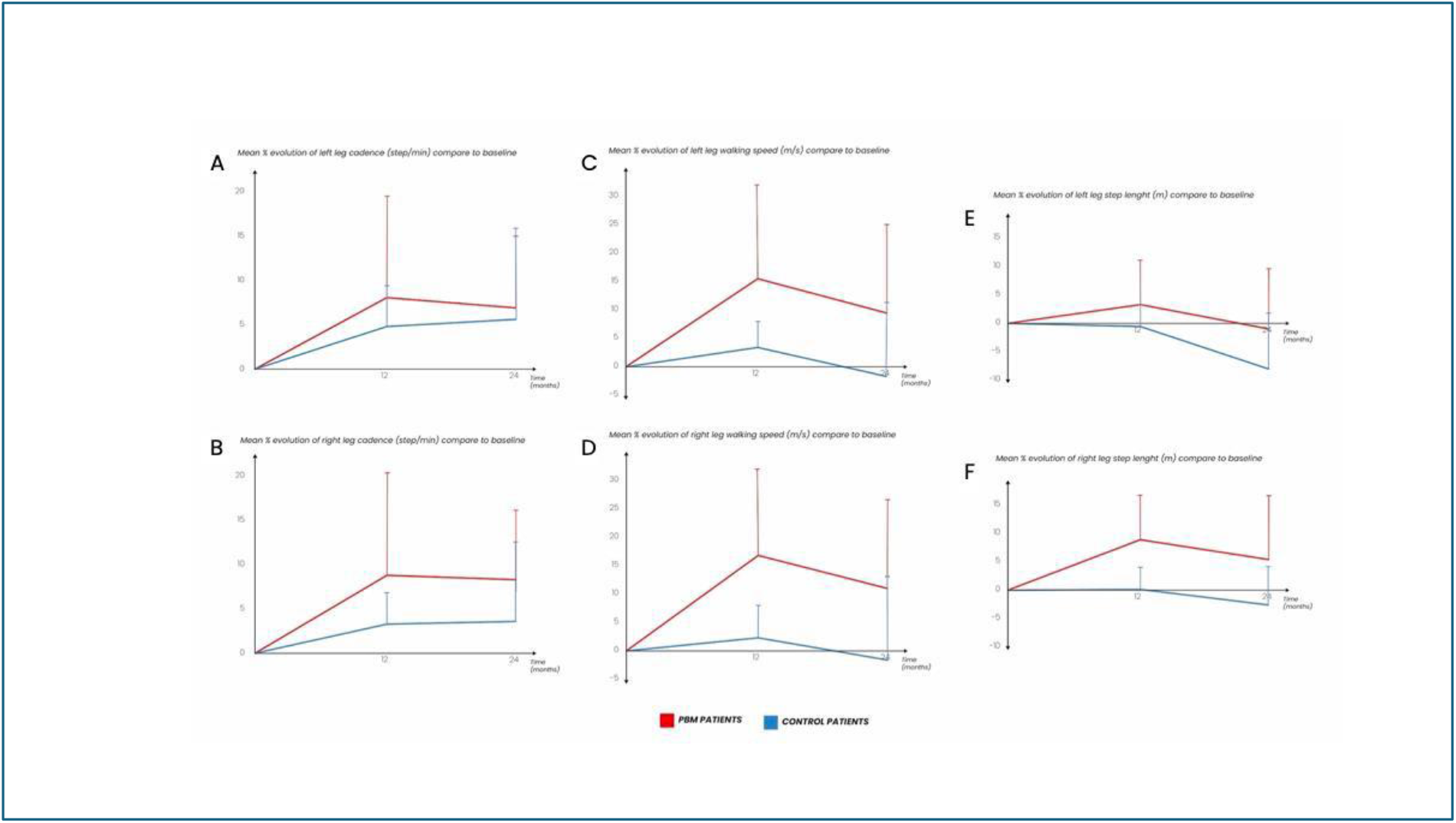
<u>The figure 4</u> shows the evolution of the spatiotemporal gait parameters obtained by a 3D gait analysis (Vicon, Biometrics). This system consisted of 8 infrared camera able to detect signals reflected by passive markers. Markers are positioned on each foot (toe, heel and ankle) to generate gait cycle parameters (Cadence, Walking Speed, Stride length, Step Time, Opposite Foot Contact…). Data acquisition started by asking the patients to stand up and walk on a distance of 5 meters and to come back during three sessions. Values are expressed by the percentage of evolution between the raw data at Month 0 and Month 12 (left panel) and Month 24 (right panel) for 3 parameters for PBM (red lines) and control group (Blue line): A and B shows the evolution of cadence (steps/ min) for left and right leg respectively. C and D show the walking Speed (in m/ sec) for left and right leg. E and F show the stride Length (m) for left and right leg.

#### Control Group

[^11^ C]-PE2I-PET at 12 months compared to baseline showed a mean (SD) bilateral decrease of [^11^C]PE2I BPND, as expected, in the caudate nucleus -16.96% (−16.3%), the nucleus accumbens -27.54% (−28.4%), and the putamen -19.52% (−11.4%), but a slight increase in the pallidum +10.74% (+39.6%).

### Adverse events

Main adverse events reported consisted in mood fluctuation (iPBM#1), substantial weight gain (4 to 5 kg) relative to baseline (all iPBM patients) and mild sleep perturbation (iPBM#2) already experienced by the patient. Importantly, patients did not feel any sensations when illumination was turned on. No abnormalities were detected in eye fundus examinations.

## DISCUSSION

In this small cohort of de novo PD patients, we show that chronic iPBM has a potential disease-modifying effect, based on convergent clinical improvement and [^11^C]-PE2I-PET data. Patients reported no major adverse effects, consistent with animal experiments already described ^10,13^. The most striking finding was that, after 12 months, iPBM induced a substantial increase in dopamine transporter levels, as measured by [^11^C]-PE2I-PET across all basal ganglia in 2 out of 3 patients. For iPBM #3, results were more dispersed depending on the basal ganglia area. In contrast, in the control group and across all basal ganglia, [^11^ C]-PE2I uptake was reduced as expected ^14^. These findings suggest that iPBM could restore dopaminergic function which supports a potential disease-modifying effect. This result is consistent with preclinical studies in several animal models of PD ^10,13^. The mechanism of action is not entirely clear, but iPBM could improve mitochondrial activity and increases ATP levels, while also inducing long-term activation of genes promoting cell survival ^15^. Dopaminergic neurons which are suffering but still alive could thus restore their production of dopamine.

Importantly, the improvements in dopamine transporter levels were associated with improvement of MDS-UPDRS motor scores. For iPBM#3, a trend for clinical improvement was observed after adjustment of the illumination protocol. These results contrast with the worsening of clinical manifestations in the control group, in line with the expected progression^16^.

Analysis of walking parameters showed an improvement of walking speed, cadence and step length for iPBM patients, whereas only cadence was improved in the control group.

The evolution of clinical and imaging data after chronic iPBM is of great importance and strengthens the hypothesis of a disease-modifying therapy through an increase in dopamine transporter levels and possibly in the numbers of dopaminergic terminals.

## Study limitations

The patient population size does not allow us to definitively conclude on a disease-modifying effect. As this was an open-label study, it is impossible to discount the possible contribution of a (partial) placebo effect in the improvement to clinical symptoms observed which is very unlikely to persist at 24 months. In addition, a persistent symptomatic effect of dopaminergic medications cannot be excluded, despite the 24-hour withdrawal of anti-parkinsonian drugs before performing clinical evaluations. However, residual effects cannot account for the improved [^11^C]PE2I-PET profiles recorded for the treated patients as it is well-established that anti-parkinsonian medications have no effect on [^11^C]PE2I-PET^17^.

## CONCLUSION

iPBM is both well tolerated and safe for use in PD patients and shows promise as a mean to modify disease progression. A larger number of patients with longer follow up is mandatory to validate the clinical efficacy of iPBM in slowing down PD progression.

## Data Availability

All data produced in the present study will be available upon reasonable request to the authors, after completion of the study

## Acknowledgements

CHU Grenoble was the promotor of this study, providing technical resources and ensuring regulatory approvals; SurgiQual Institute provided regulatory requirements. The authors wish to thank all collaborators from Clinatec for their help, especially Caroline Sandre-Ballester, Marion Coquand-Gandit, Stéphane Pezzani, Nelly Richard, Lilia Langar, Madjid Hihi, and Virginie Brun. We also thank Boston Scientific and Mike Moffit, the Edmond J Safra Foundation and the “Fonds Clinatec” for their support. We are very grateful to all those who were involved in the initiation and development of the methods and the early trials that led to the development of the study protocol.

## Notes

### Competing Interest Statement

SC is consultant for Medtronic and Boston Scientific
ST has received consultant fees from Abbvie, Boston Scientific, Medtronic and payements for lectures from MERZ, Movement Disorders Society, NHC, Aguettant

### Clinical Trial

NCT04261569

### Funding Statement

This study was funded by Boston Scientific and the Edmond J Safra Foundation

### Author Declarations

The details of the Institutional Review Board (IRB) or oversight body that approved or provided exemption for the research described in this manuscript are outlined below: All analyses conducted as part of the present study were approved by the Comite de Protection des Personnes Phones Alpes Auvergne (2019-A02097-50) April 2020 and received additional authorization from the French National Agency for the Safety of Medicines and Health Products (ANSM) in January 2020. The treatment and handling of data adhered to all applicable ethical regulations.

## REFERENCES

1. Pagano G, Taylor KI, Anzures-Cabrera J, et al. Trial of Prasinezumab in Early-Stage Parkinson’s Disease. New England Journal of Medicine. 2022;387(5):421–432. doi:10.1056/nejmoa2202867

2. Lang AE, Siderowf AD, Macklin EA, et al. Trial of Cinpanemab in Early Parkinson’s Disease. New England Journal of Medicine. 2022;387(5):408–420. doi:10.1056/nejmoa2203395

3. Devos D, Labreuche J, Rascol O, et al. Trial of Deferiprone in Parkinson’s Disease. New England Journal of Medicine. 2022;387(22):2045–2055. doi:10.1056/nejmoa2209254

4. Meissner WG, Remy P, Giordana C, et al. Trial of Lixisenatide in Early Parkinson’s Disease. New England Journal of Medicine. 2024;390(13):1176–1185. doi:10.1056/nejmoa2312323

5. González-Rodríguez P, Zampese E, Stout KA, et al. Disruption of mitochondrial complex I induces progressive parkinsonism. Nature. 2021;599(7886):650–656. doi:10.1038/s41586-021-04059-0

6. Foo ASC, Soong TW, Yeo TT, Lim KL. Mitochondrial Dysfunction and Parkinson’s Disease—Near-Infrared Photobiomodulation as a Potential Therapeutic Strategy. Front Aging Neurosci. 2020;12. doi:10.3389/fnagi.2020.00089

7. Ramsay RR, Salach JI, Singer TP. Uptake of the neurotoxin 1-methyl-4-phenylpyridine (MPP+) by mitochondria and its relation to the inhibition of the mitochondrial oxidation of NAD+-linked substrates by MPP+. Biochem Biophys Res Commun. 1986;134(2):743–748. doi:10.1016/S0006-291X(86)80483-1

8. Michel PP, Hirsch EC, Hunot S. Understanding Dopaminergic Cell Death Pathways in Parkinson Disease. Neuron. 2016;90(4):675–691. doi:10.1016/j.neuron.2016.03.038

9. Hamblin MR, Liebert A. Photobiomodulation Therapy Mechanisms Beyond Cytochrome c Oxidase. Photobiomodul Photomed Laser Surg. 2022;40(2):75–77. doi:10.1089/photob.2021.0119

10. Darlot F, Moro C, El Massri N, et al. Near-infrared light is neuroprotective in a monkey model of Parkinson disease. Ann Neurol. 2016;79(1):59–75. doi:10.1002/ana.24542

11. Reinhart F, Massri N El, Torres N, et al. The behavioural and neuroprotective outcomes when 670nm and 810nm near infrared light are applied together in MPTP-treated mice. Neurosci Res. 2017;117:42–47. doi:10.1016/j.neures.2016.11.006

12. Moro C, Massri N El, Torres N, et al. Photobiomodulation inside the brain: a novel method of applying near-infrared light intracranially and its impact on dopaminergic cell survival in MPTP-treated mice: Laboratory investigation. Journal of Neurosurgery JNS. 2014;120(3):670–683. doi:10.3171/2013.9.JNS13423

13. Moro C, Torres N, Arvanitakis K, et al. No evidence for toxicity after long-term photobiomodulation in normal non-human primates. Exp Brain Res. 2017;235(10):3081–3092. doi:10.1007/s00221-017-5048-7

14. Li W, Lao-Kaim NP, Roussakis AA, et al. 11C-PE2I and 18F-Dopa PET for assessing progression rate in Parkinson’s: A longitudinal study. Movement Disorders. 2018;33(1):117–127. doi:10.1002/mds.27183

15. Hamblin MR. Photobiomodulation or low-level laser therapy. J Biophotonics. 2016;9(11-12):1122-1124. doi:10.1002/jbio.201670113

16. Holden SK, Finseth T, Sillau SH, Berman BD. Progression of MDS-UPDRS Scores Over Five Years in De Novo Parkinson Disease from the Parkinson’s Progression Markers Initiative Cohort. Mov Disord Clin Pract. 2018;5(1):47–53. doi:10.1002/mdc3.12553

17. Bang JI, Jung IS, Song YS, et al. PET imaging of dopamine transporters with [18F]FE-PE2I: Effects of anti-Parkinsonian drugs. Nucl Med Biol. 2016;43(2):158–164. doi:10.1016/j.nucmedbio.2015.11.002

18. Chabardès S, Carron R, Seigneuret E, et al. Endoventricular Deep Brain Stimulation of the Third Ventricle: Proof of Concept and Application to Cluster Headache. Neurosurgery. 2016;79(6). doi:10.1227/NEU.0000000000001260

19. Tomlinson CL, Stowe R, Patel S, Rick C, Gray R, Clarke CE. Systematic review of levodopa dose equivalency reporting in Parkinson’s disease. Movement Disorders. 2010;25(15):2649–2653. doi:10.1002/mds.23429

